# Exposure to Prenatal Social Disadvantage and Maternal Psychosocial Stress: Relationships to Neonatal White Matter Connectivity

**DOI:** 10.1101/2022.05.10.22274924

**Authors:** Rachel E. Lean, Christopher D. Smyser, Rebecca G. Brady, Regina L. Triplett, Sydney Kaplan, Jeanette K. Kenley, Joshua S. Shimony, Tara A. Smyser, J. Phillip Miller, Deanna M. Barch, Joan L. Luby, Barbara B. Warner, Cynthia E. Rogers

**Author notes:** **Corresponding Author:** Rachel E. Lean. 4444 Forest Park Ave, Campus Box 8514, St. Louis, MO 63110. **Competing Interest Statement:** The authors have no real or perceived competing interests to disclose.

## Abstract

Childhood exposure to poverty and related psychosocial stressors are associated with altered microstructure in fronto-limbic pathways evident at age 8-10 years. How early in neurodevelopment these associations can be detected remains unclear. In this longitudinal study, 399 mothers were oversampled for low income and completed social experience and background measures during pregnancy. Measures were analyzed with structural equation analysis resulting in two latent constructs: Social Disadvantage (education, insurance status, income-to-needs ratio [INR], neighborhood deprivation, nutrition) and Psychosocial Stress (depression, stress, life events, racial discrimination). At birth, 289 healthy term-born neonates underwent a diffusion MRI (dMRI) scan. Mean diffusivity (MD) and fractional anisotropy (FA) were measured for the dorsal and inferior cingulum bundle (CB), uncinate, and fornix using probabilistic tractography in FSL. Social Disadvantage and Psychosocial Stress were fitted to dMRI parameters using regression models adjusted for infant postmenstrual age at scan and sex. Social Disadvantage, but not Psychosocial Stress, was independently associated with lower MD in the bilateral inferior CB and left uncinate, right fornix, and lower MD and higher FA in the right dorsal CB. Results persisted after accounting for maternal medical risk in pregnancy and prenatal drug exposure. In moderation analysis, Psychosocial Stress was associated with lower MD in the left inferior CB among the lower-to-higher SES (INR ≥200%) group, but not the extremely low SES (INR <200%) group. Increasing access to social welfare programs that reduce the burden of poverty and related psychosocial stressors may be an important target to protect fetal brain development in fronto-limbic pathways.

## Introduction

Early childhood is a sensitive period of brain development that is highly susceptible to exposure to poverty (1). Various forms of childhood social disadvantage have been associated with lasting reductions in prefrontal, amygdala and hippocampal volumes (2). However, less is known about these effects on white matter development more specifically, including their regional specificity and timing. The cingulum bundle (CB), uncinate, and fornix are key white matter tracts that connect fronto-limbic brain regions and are similarly altered by poverty (3, 4). Studies using diffusion magnetic resonance imaging (dMRI) have shown that childhood social disadvantage is related to lower fractional anisotropy (FA) in the uncinate, fornix, and CB in children age 8-10 years (4) and adults (3).One study has focused on perinatal social disadvantage in preterm and term-born neonates, linking family disadvantage with lower FA in the uncinate at term-equivalent age (5). This study, however, focused on family-level factors including caregiver education, employment status, occupation, primary language spoken, age at child delivery, and family structure. In addition to family-level factors, impacts of the broader social environment including neighborhood socioeconomic status (SES) have also been related to brain development in childhood (6).

Exploring prenatal and early postnatal effects may be particularly important due to the rapid proliferation of immature oligodendrocyte cells starting from 28 weeks gestation, followed by myelination of white matter fibers in the first weeks of life which progresses in an orderly, regionally-specific, anterior-posterior manner (7–9). Thus, prenatal white matter development is a critical period of heightened neuronal plasticity which may be disrupted by early life adversity *in utero* (10). Yet, little is known about the impact of social disadvantage, as a form of adversity, on prenatal white matter development in humans. Preclinical work in rodents suggests that resource deprivation and exposure to other related stressors produces pro-inflammatory cytokines and inhibits the expression of proteins responsible for axonal differentiation and myelination, resulting in fewer mature, myelin producing oligodendrocytes in developing white matter (11–13). Exposure to poverty and related stressors may also exacerbate reactive oxygen species, causing both direct injury to and resulting apoptosis of oligodendrocyte precursors (14) and arrest of oligodendrocyte differentiation without cell death (15). Taken together, these findings suggest that *in utero* exposure to social disadvantage may impair the microstructural maturation of white matter in developing humans during a period of high brain plasticity.

While childhood exposure to poverty alters white matter connectivity (3, 4), poverty co-occurs with other forms of adversity such as exposure to parental psychopathology and stressful life events (16). One pathway by which poverty may shape offspring brain development includes maternal psychosocial stress. To date, only a handful of studies have examined maternal psychosocial stress in pregnancy and neonatal white matter connectivity (17–20). Maternal anxiety in the third trimester has been shown to predict lower FA in the CB, fornix, and uncinate in neonates (18, 21), and higher mean diffusivity (MD) in prefrontal white matter at age 1 month (19); white matter regions also impacted by poverty (3, 4). Maternal perinatal depression has been related to higher MD in frontal white matter (19) and fewer white matter fibers projecting from the amygdala to the pre-frontal cortex in neonates (20). While these findings highlight the role of maternal psychosocial stress, the extent to which social disadvantage may have a dissociable impact on white matter connectivity at birth is unclear. Prior studies of maternal psychosocial stress have typically examined social disadvantage as a confound in adjusted analyses (17, 18, 20). However, treating social disadvantage as a confound, rather than as an independent variable of interest, limits the understanding of its direct contribution to early brain development.

This study focuses on 289 healthy term-born neonates with brain neuroimaging acquired at birth. Mothers were identified in the first trimester of pregnancy and oversampled for poverty. As described in Luby et al. (22), observed variables collected during pregnancy were used to define two latent constructs: Social Disadvantage (education, health insurance, Income-to-Needs Ratio [INR], neighborhood poverty, nutrition) and Psychosocial Stress (depression, stress, life events, racial discrimination; see also Supplementary Information). This study examines prenatal exposure to Social Disadvantage and Psychosocial Stress in relation to white matter connectivity at birth. We hypothesized that Social Disadvantage and Psychosocial Stress would be independently associated with CB, uncinate, and fornix microstructure (Figure 1). These fronto-limbic tracts were selected due to their vulnerability to early life adversity (3, 4) and importance for socio-emotional development (23, 24). We also included the corpus callosum (CC) as a negative control tract because it is a large anterior-posterior tract in proximity to the CB with branching fibers, but it does not connect fronto-limbic brain regions. As postnatal exposure to maternal stress mediates the effect of poverty on macro-structural brain development (25), we hypothesized that Psychosocial Stress would explain more variance in white matter connectivity than Social Disadvantage. We also examined whether associations between Psychosocial Stress and neonatal white matter varied as a function of family SES group, defined as INR below or at/above 200% of the national poverty threshold (26). Supplementary aims included examining maternal medical risk (MMR) in pregnancy and prenatal drug exposure (cannabis and tobacco) as confounding factors.

**Figure 1.**
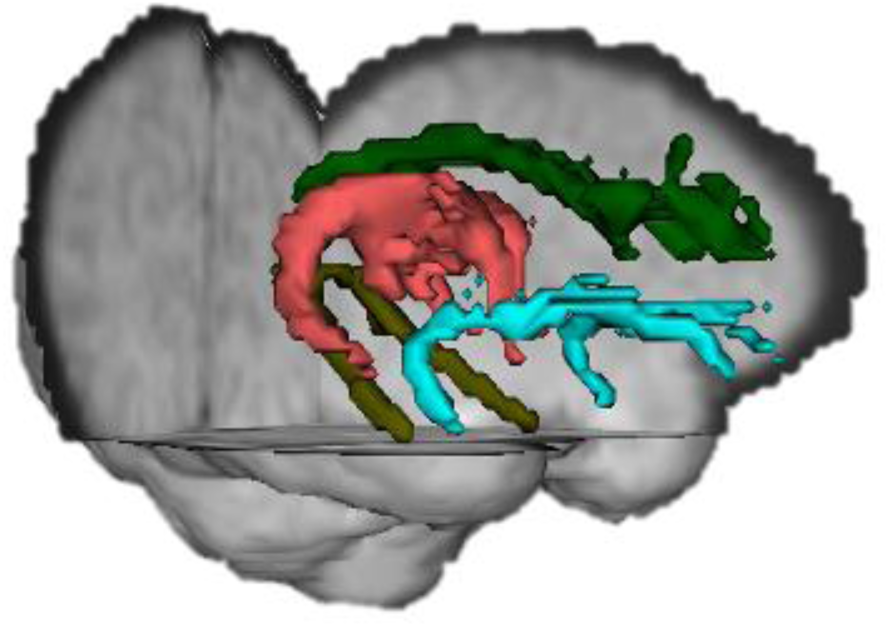
Key fronto-limbic white matter tracts in a representative healthy, term-born neonate. Tracts of interest include the dorsal (green) and inferior (gold) cingulum bundle, fornix (pink), and uncinate (blue).

## Materials and Methods

### Sample

This study draws data from a longitudinal study of infants (born 2017–2020) and their mothers (recruited 09/2017–02/2020) participating in the Early Life Adversity and Biological Embedding (eLABE) Study (22). Pregnant women were identified from the March of Dimes Prematurity Research Center at Washington University in St. Louis. eLABE study exclusion criteria spanned multiple gestations, infections known to cause congenital disease (*e*.*g*., syphilis), and alcohol or drug use other than tobacco and cannabis. The eLABE study recruited 395 pregnant women (declined participation *n*=268) and their 399 singleton infants (4 mothers had 2 singleton births during recruitment). Women were over-sampled from an antenatal clinic serving low-income women to enrich the sample for exposure to poverty.

### Procedure

Mothers completed surveys in each trimester and at delivery to assess social background, mental health, and life experiences. Medical data were collected from surveys and chart review. In the first weeks of life, 385 non-sedated neonates underwent an MRI scan on a Siemens Prisma 3T scanner with a 64-channel head coil (Figure S1, Supplementary Information). After feeding, neonates were swaddled, positioned in a head-stabilizing vacuum fix wrap, fitted with noise protection gear, and placed in the head coil on foam padding to decrease motion. MRI scans were not performed for 14 neonates due to the COVID-19 Pandemic (March, 2020). dMRI data was obtained for 365/385 neonates (no DTI sequence collected *n*=3, required frames not collected *n*=4, sequence collected in one direction *n*=8, artifact *n*=5). Seventy-six neonates were excluded from analyses due to preterm birth (<37 weeks gestation *n*=53), low birthweight (<2000 grams *n*=1), Neonatal Intensive Care Unit admission >7 days (*n*=8), high-grade brain injury (*n*=14). Thus, 289 healthy term-born neonates were included in analyses (PMA at scan *m*=42 weeks, SD=1.3, range 38-45) (Table 1). Study procedures were approved by the Institutional Review Board. Written informed consent was obtained from all mothers.

**Table 1.**
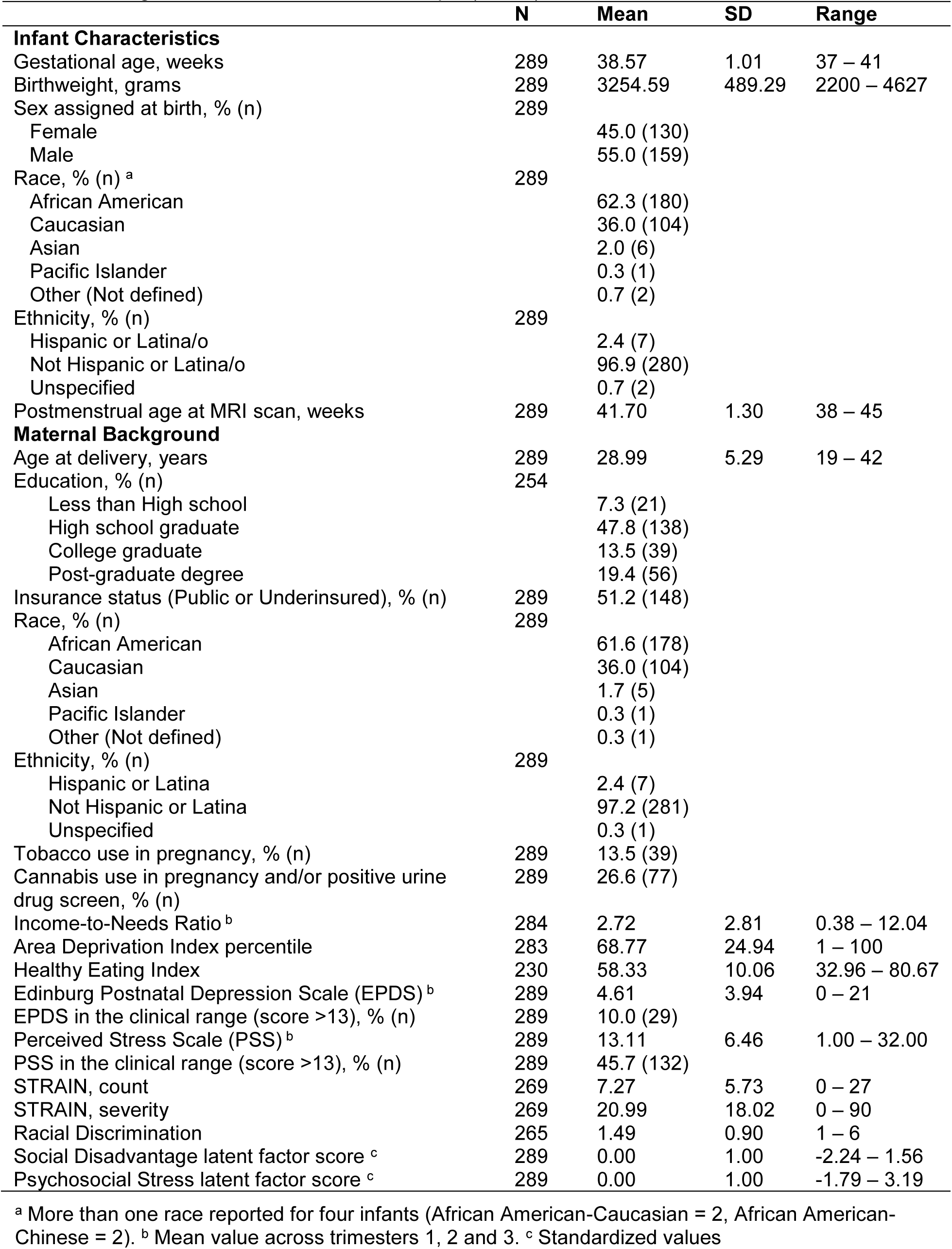
Background characteristics of the sample (*n*=289).

### Measures

#### Social Disadvantage

Maternal education level and health insurance status were collected in trimester 1. Mothers reported household income and persons in the home to calculate Income-to-Needs Ratio (INR) (43) in each trimester. Home addresses were collected at delivery to obtain Area Deprivation Index (ADI) percentiles (49) to asses neighborhood SES, housing quality, and access to necessities using Census block data. Prior work in the eLABE cohort has shown that while 26% of mothers changed addresses during pregnancy, there was no significant change in block group (50). Mothers completed the Healthy Eating Index-2015 (51) in the third trimester or at delivery to assess nutrition.

#### Psychosocial Stress

In each trimester, mothers completed the Edinburgh Postnatal Depression Scale (EPDS) (52) and the Cohen Perceived Stress Scale (53) to assess depression symptoms and perceived stress. Stressful/traumatic life events (count and severity) was assessed with the Stress and Adversity Inventory for Adults (STRAIN) (54), collected at the neonatal MRI scan (*n*=183) or at the one year follow-up (*n*=86). There was no difference in STRAIN count (*d*=0.05, *p*=.71) or severity (*d*=0.07, *p*=.58) between mothers who completed the STRAIN at the neonatal MRI scan or at follow-up. The Everyday Discrimination Survey (55) measured racial discrimination, collected at the neonatal MRI scan.

#### Latent Factors

In Luby et al. (22), observed variables were analyzed for all eLABE mothers using structural equation analysis, resulting in the latent factors Social Disadvantage and Psychosocial Stress (Figure S2, Supplementary Information). Social Disadvantage was similar for subjects excluded or included in analyses (*d*=0.03, *p*=.76) but there was a difference in Psychosocial Stress (*d*=0.36, *p*=.001).

#### White Matter Microstructure

Neonatal dMRI scans were acquired as two five-minute runs using MB4, TR/TE=2500/79.4 ms, (1.75 mm)^3^ voxels, with whole brain coverage (80 slices), 108 b values sampled on 3 shells of b=500-2500 s/mm^2^ and 7 b=0 images interspersed throughout each run with phase encoding direction reversal (anterior → posterior and posterior → anterior) for susceptibility- and eddy-current distortion correction (56). dMRI parameters were extracted from the dorsal and inferior CB, fornix, and uncinate (Figure 1) using probabilistic tractography in FSL Version 5.0.9. Anatomically landmarked seeds were placed by two highly trained raters (inter-rater coefficients: 0.80–0.98 for MD and 0.73–0.92 for FA).

### Data Analysis

Continuous variables were examined for distributions and outliers. Distributions were found to approximate normal. Four MD values (1 each for the left uncinate, right uncinate, right inferior CB, and right fornix) were extreme outliers (>3 SD) and removed from analysis. Curve Estimation was used to explore fit of associations between latent factors and dMRI parameters, with linear models providing the best overall fit. Social Disadvantage and Psychosocial Stress were examined separately in relation to dMRI parameters using regression models adjusted for infant PMA at scan and sex (male=1, female=2). RD and AD were also examined for tracts with significant MD results. Next, Social Disadvantage and Psychosocial Stress were fitted together in hierarchical, multivariable linear regression models. Standardized coefficients, standard errors, and R^2^ values are reported. In step 1 of the regression, infant PMA at scan and sex were entered as covariates. Social Disadvantage and Psychosocial Stress were entered in step 2, with R^2^ change statistics reported. Bivariate and multivariable analyses were multiple comparison corrected using the Benjamini-Hochberg False Discovery Rate procedure (8 corrections [number of tracts] for each dMRI parameter). Moderation analysis was used to test interactions between Psychosocial Stress and family SES groups on white matter connectivity. Supplementary regression models were used to examine MMR during pregnancy and prenatal drug (cannabis and tobacco) exposure as founding factors (Supplementary Information).

### Data Sharing Plans

Data were collected with consent forms that allow data sharing. De-identified data is being deposited in the NIMH Data Archive and will be available after the conclusion of the eLABE 3-year wave.

## Results

Sample characteristics are shown in Table 1. First, prenatal exposure to Social Disadvantage and Psychosocial Stress were examined separately in relation to dMRI parameters, adjusted for infant PMA at scan and sex (Tables S1, S2; Supplementary Information). Then, to delineate the conditional contribution of each latent construct when the other was included in the model, Social Disadvantage and Psychosocial Stress were fitted together in hierarchical regression models (results from step 2 of the regression models are summarized in Table 2). Results were multiple comparison corrected using Benjamini-Hochberg False Discovery Rate. Full results (Tables S3, S4) show that infant PMA at scan explained 15–31% of the variance in MD and 9–26% of the variance in FA across tracts.

**Table 2.** Summary of hierarchical regression models linking infant characteristics and early life adversity constructs with neonatal Mean Diffusivity (MD) Fractional Anisotropy (FA) (*n*=289).

|  | MD |  |  |  | FA |  |  |  |
| --- | --- | --- | --- | --- | --- | --- | --- | --- |
| | $\beta$ | SE | $p$ | $q$ | $\beta$ | SE | $p$ | $q$ |
| **R Dorsal Cingulum** | $R^2=.25, p<.001$ | | | | $R^2=.13, p<.001$ | | | |
| Sex | -.022 | .051 | .66 | .87 | -.111 | .056 | .05 | .13 |
| PMA at scan | -.504 | .052 | <.001 | <.001 | .321 | .057 | <.001 | <.001 |
| Social Disadvantage | -.168 | .056 | .003 | .008 | .179 | .061 | .003 | .02 |
| Psychosocial Stress | .015 | .055 | .79 | .94 | -.068 | .060 | .26 | .57 |
| **L Dorsal Cingulum** | $R^2=.22, p<.001$ | | | | $R^2=.13, p<.001$ | | | |
| Sex | -.022 | .052 | .67 | .87 | -.160 | .056 | .004 | .03 |
| PMA at scan | -.471 | .053 | <.001 | <.001 | .301 | .057 | <.001 | <.001 |
| Social Disadvantage | -.103 | .057 | .07 | .07 | .135 | .061 | .03 | .07 |
| Psychosocial Stress | -.059 | .056 | .29 | .94 | -.025 | .060 | .68 | .83 |
| **R Inferior Cingulum** | $R^2=.29, p<.001$ | | | | $R^2=.11, p<.001$ | | | |
| Sex | -.104 | .051 | .04 | .16 | .057 | .056 | .31 | .38 |
| PMA at scan | -.516 | .052 | <.001 | <.001 | .342 | .057 | <.001 | <.001 |
| Social Disadvantage | -.246 | .055 | <.001 | <.001 | .047 | .061 | .44 | .71 |
| Psychosocial Stress | .004 | .055 | .94 | .94 | .021 | .060 | .72 | .83 |
| **L Inferior Cingulum** | $R^2=.36, p<.001$ | | | | $R^2=.11, p<.001$ | | | |
| Sex | -.109 | .048 | .02 | .16 | -.055 | .057 | .34 | .38 |
| PMA at scan | -.588 | .049 | <.001 | <.001 | .305 | .058 | <.001 | <.001 |
| Social Disadvantage | -.214 | .052 | <.001 | <.001 | -.006 | .062 | .92 | .97 |
| Psychosocial Stress | -.050 | .052 | .33 | .94 | .101 | .061 | .10 | .57 |
| **R Uncinate** | $R^2=.21, p<.001$ | | | | $R^2=.26, p<.001$ | | | |
| Sex | -.056 | .053 | .30 | .69 | -.114 | .051 | .03 | .11 |
| PMA at scan | -.464 | .054 | <.001 | <.001 | .492 | .052 | <.001 | <.001 |
| Social Disadvantage | -.110 | .058 | .06 | .07 | .002 | .056 | .97 | .97 |
| Psychosocial Stress | -.012 | .057 | .84 | .94 | -.011 | .055 | .84 | .84 |
| **L Uncinate** | $R^2=.16, p<.001$ | | | | $R^2=.19, p<.001$ | | | |
| Sex | -.051 | .055 | .35 | .69 | -.004 | .053 | .94 | .94 |
| PMA at scan | -.396 | .056 | <.001 | <.001 | .429 | .054 | <.001 | <.001 |
| Social Disadvantage | -.169 | .060 | .005 | .01 | .016 | .058 | .78 | .97 |
| Psychosocial Stress | -.006 | .059 | .91 | .94 | .082 | .058 | .16 | .57 |
| **R Fornix** | $R^2=.32, p<.001$ | | | | $R^2=.11, p<.001$ | | | |
| Sex | .008 | .049 | .87 | .87 | -.084 | .057 | .14 | .22 |
| PMA at scan | -.569 | .050 | <.001 | <.001 | .311 | .058 | <.001 | <.001 |
| Social Disadvantage | -.141 | .054 | .009 | .01 | .140 | .062 | .03 | .07 |
| Psychosocial Stress | .029 | .053 | .59 | .94 | -.066 | .061 | .28 | .57 |
| **L Fornix** | $R^2=.31, p<.001$ | | | | $R^2=.10, p<.001$ | | | |
| Sex | .010 | .049 | .84 | .87 | -.097 | .057 | .09 | .18 |
| PMA at scan | -.564 | .050 | <.001 | <.001 | .278 | .058 | <.001 | <.001 |
| Social Disadvantage | -.096 | .054 | .07 | .07 | .107 | .062 | .09 | .17 |
| Psychosocial Stress | .013 | .053 | .80 | .94 | .039 | .061 | .52 | .83 |

*Note.* R, Right; L, Left; SE, Standard Error; $q$ , significance value adjusted for multiple comparisons using Benjamini-Hochberg False Discovery Rate procedure; PMA, postmenstrual age at scan.

### Social Disadvantage

As shown in Table S1 (Supplementary Information), Social Disadvantage was related to lower MD in the bilateral dorsal and inferior CB, bilateral uncinate, and right fornix. Results persisted after multiple comparison correction. MD findings for the bilateral dorsal and inferior CB, left uncinate, and right fornix were due to lower radial diffusivity (RD), with the bilateral inferior CB and left uncinate also showing lower axial diffusivity (AD). Associations between Social Disadvantage and higher FA in the bilateral dorsal CB and bilateral fornix did not survive multiple comparison correction.

### Psychosocial Stress

As shown in Table S2 (Supplementary Information), Psychosocial Stress was associated with lower MD in the left inferior CB, driven by reduced RD. This result persisted after multiple comparison correction. There were no associations for the uncinate or fornix.

### Multivariable Analyses of Social Disadvantage and Psychosocial Stress

When Psychosocial Stress and Social Disadvantage were fitted together in step 2 of the regression analyses (Table 2), Social Disadvantage continued to explain significant independent variance in MD for the right dorsal CB, bilateral inferior CB, left uncinate, and right fornix as well as higher FA in the right dorsal CB. Adding Social Disadvantage to step 2 of the multivariable model explained an additional 2 – 6% of the variance in MD and FA across white matter tracts (Table S3, S4; Supplementary Information). Importantly, the association between Psychosocial Stress and left inferior CB MD was attenuated after accounting for Social Disadvantage (Table 2). There were no unique associations between Social Disadvantage or Psychosocial Stress and FA in the uncinate or fornix after multiple comparison correction (Table 2). A similar pattern of null results was also observed for the CC as a negative control tract (Table S5).

### Psychosocial Stress in Extremely Low and Lower-to-Higher SES Groups

Given the bivariate association between Psychosocial Stress and left inferior CB MD prior to accounting for Social Disadvantage (Table S2, Supplementary Information), we examined whether the strength of the association between Psychosocial Stress and the inferior CB varied as a function of family SES. INR was used to dichotomize the sample into extremely low and lower-to-higher family SES groups (defined as mean INR below or at/above 200% of the national poverty threshold (26), respectively) because INR loaded most heavily on the Social Disadvantage factor (22). Moderation analysis (Table S6) showed that there was an interaction between Psychosocial Stress and family SES group on left inferior CB MD (B= - .31, *p*=.008), such that the association between Psychosocial Stress and left inferior CB MD was stronger in the lower-to-higher SES group than in the extremely low SES group (Figure 2). Within the lower-to-higher SES group, the effect of Psychosocial Stress on the inferior CB persisted after also accounting for individual differences in Social Disadvantage factor scores (Table S7).

**Figure 2.**
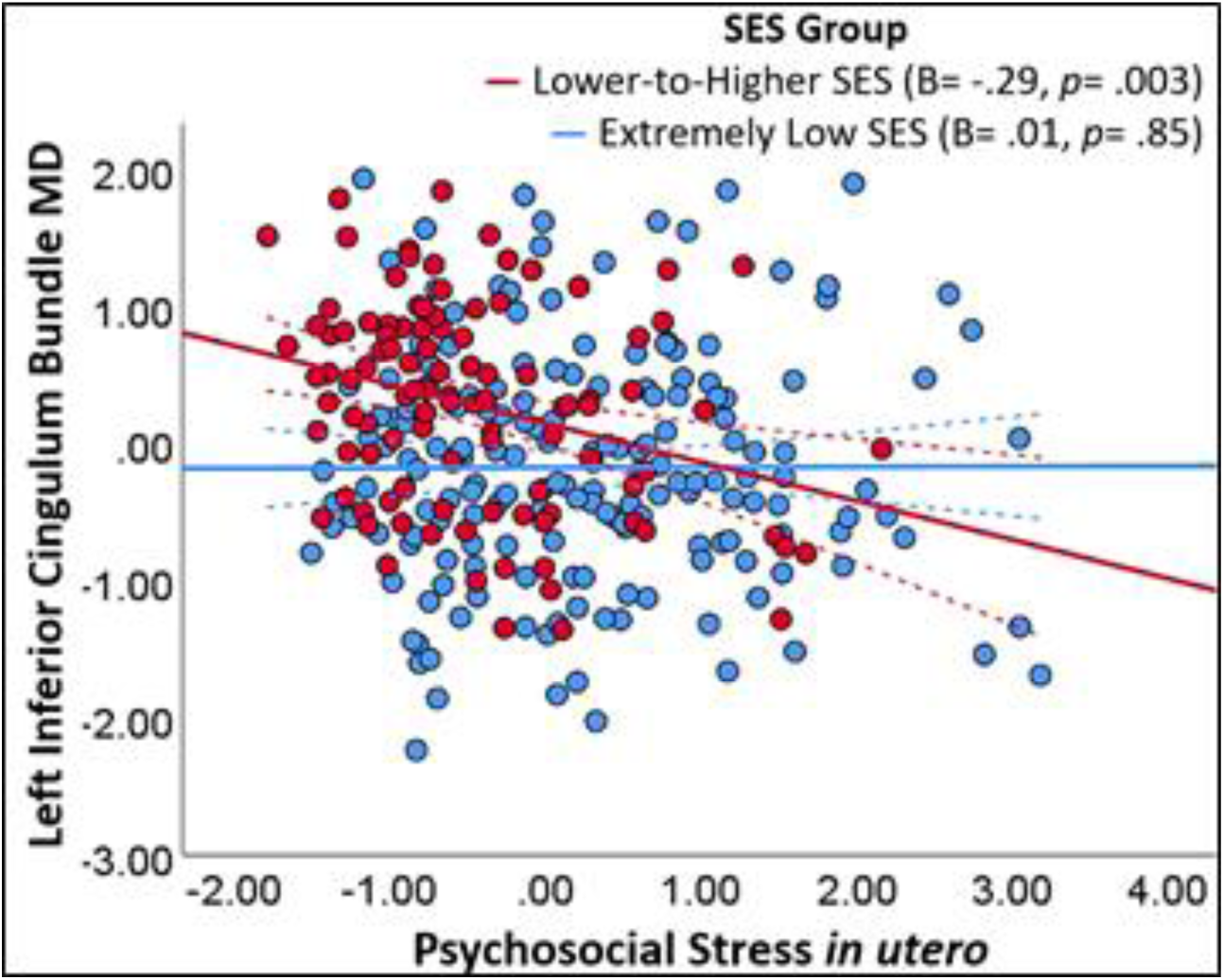
The interaction between Psychosocial Stress *in utero* and family socioeconomic status (SES) group on the left inferior cingulum bundle mean diffusivity (MD) at birth (*n*=284). Here, MD values are shown as standardized residuals adjusted for infant PMA at scan. Solid lines represent the slope of each SES group and dashed lines represent 95% confidence intervals.

### Confounding Factors

As described in Supplementary Information, MMR was only correlated with higher MD in the left dorsal CB (*p*=.03). Prenatal exposure to cannabis was related to lower MD in the left inferior CB (*p*=.04) and higher FA in the left fornix (*p*=.02). Tobacco exposure was not significant. As reported in Tables S8 and S10 (Supplementary Information), MMR and cannabis exposure were not significant after accounting for infant PMA at scan and, therefore, did not alter the main conclusions of this study.

## Discussion

In healthy term-born neonates, prenatal exposure to Social Disadvantage was independently related to lower MD in the bilateral inferior CB, left uncinate, and right fornix, and lower MD and higher FA in the right dorsal CB after accounting for covariate factors and maternal Psychosocial Stress. While maternal Psychosocial Stress was correlated with lower MD in the left inferior CB, this finding did not persist after accounting for Social Disadvantage. However, moderation analysis showed that Psychosocial Stress was more strongly associated with lower MD in the left inferior CB among lower-to-higher SES dyads. Key study findings persisted after accounting for MMR and prenatal exposure to cannabis.

Consistent with Thompson et al. (5), Social Disadvantage *in utero* was associated with aberrant white matter connectivity at birth. Thompson *et al*. found alterations mainly in the uncinate, and we found effects in the CB, uncinate, and fornix, but not the CC. Our results suggest that despite differences in timing of myelination (27), prenatal exposure to Social Disadvantage is associated with variability in white matter connectivity across fronto-limbic tracts of interest. Findings of lower MD or higher FA could reflect aberrant reductions in fiber branching or atypical pruning of white matter fibers, making the underlying dMRI tensors appear more directional (23, 28). Indeed, the CB and uncinate contain multiple branching fibers (28, 29), and the fornix is a highly angular tract with projections into the prefrontal cortex (30). Lower MD or higher FA early in development may also be a marker of hyper-maturation (31, 32). Preclinical studies suggest that deprivation and stress drive precocious oligodendrocyte differentiation (33) and premature myelination (34) in frontal and temporal brain regions, and human studies have shown that frontal and temporal brain regions contain projections from the CB, uncinate and/or fornix (28, 29).The hyper-maturation hypothesis is particularly relevant for the CB and uncinate, given that accelerated amygdala-frontal connectivity has been linked with the over-activation of the hypothalamic-pituitary-adrenal axis and glucocorticoid expression in the context of early life adversity (32, 35). Clinically, higher neonatal and infant FA in the CB and uncinate has been shown to predict socio-emotional problems and Autism Spectrum Disorder (23, 24). Taking these findings together, social disadvantage *in utero* appears to be an important antecedent of aberrant microstructural development in key white matter tracts implicated in the development of socio-emotional impairments.

Multiple pathophysiological and environmental mechanisms likely play a role in linking poverty with altered white matter development. Because maternal obesity, pre-eclampsia, and asthma are more common in the setting of poverty (36, 37) and have been linked to micronutrient deficiencies, vascular insufficiency and/or oxidative stress likely to impair mitochondria-rich pre-oligodendrocytes in offspring (38, 39), we considered maternal medical risk (MMR) in pregnancy. MMR was related to altered microstructure in the left CB, but this result did not persist after adjusting for infant PMA at scan which captures expected age-related differences in neonatal white matter development (7–9). As we focused on healthy term-born neonates, mothers were also generally healthy. In addition, disadvantaged areas are more likely to have poorer quality housing, lack green spaces, and have greater lead toxicity and air pollution (40). Exposure to lead and air pollution *in utero* may disrupt highly sensitive oligodendrocyte differentiation, proliferation, and subsequent myelination (13–15). Lead and air pollution should be examined in future work focusing on the mechanisms linking poverty with fetal white matter development.

This study also examined prenatal exposure to Psychosocial Stress, which has been related to altered white matter connectivity at birth (17–21) and is one potential pathway by which poverty may influence macro-structural brain development in childhood (25). As such, we hypothesized that Psychosocial Stress would explain more variance in neonatal white matter connectivity than Social Disadvantage. While Psychosocial Stress was correlated with MD in the left inferior CB, this finding was accounted for or subsumed by Social Disadvantage in multivariable analyses. Prior work focusing on maternal anxiety/depression has typically controlled for SES using narrow markers like income or education (17, 18, 20). We broadly assessed social disadvantage spanning demographics, family income, neighborhood poverty, and nutrition. Compared to narrow markers of SES, multifactorial indices of social disadvantage are thought to capture general inequality, reflect the cumulative effects of multiple risk exposure, and/or reduce measurement error (41, 42). Thus, the presence of social disadvantage, a global and robust risk factor, may have a pervasive and/or more detectable impact on white matter development.

However, moderation analysis provided evidence of an interaction between Psychosocial Stress and family SES group, such that the association between Psychosocial Stress and the inferior CB was stronger in the lower-to-higher SES group than in the extremely low SES group. This suggests that maternal psychosocial stress may be more observable in the setting of less severe social disadvantage. Additionally, mothers in the lower-to-higher SES group are likely more demographically comparable to other samples in which associations between psychosocial stress and neonatal white matter development have been found (17, 19) and where psychosocial stress may be arising for reasons other than social disadvantage. We note that while we used a similar cut-point to define the lower-to-higher SES group as other large prospective studies (26), 200% of the poverty threshold is still relatively disadvantaged (*e*.*g*., income of $53,000 to support a four person household) (43). Nonetheless, the inferior CB is thought to be susceptible to psychosocial stress because it connects the amygdala with the hippocampus (28, 44). These structures are rich with glucocorticoid receptors that are highly sensitive to stress, with downstream effects on the HPA axis and production of cortisol that, in turn, alter neural plasticity (10, 44, 45).

Study strengths included the prospective study design beginning *in utero*, oversampling for exposure to poverty, broad assessment of Social Disadvantage and Psychosocial Stress, high-quality dMRI data, and a large sample compared to prior prenatal adversity studies of ≤100 term-born infants (18–21). Study limitations include the use of a self-report questionnaire to assess maternal depression. The EPDS is used in clinical and research settings and shows validity for Major Depressive Disorder (46), but mothers may have under-reported symptoms. In our sample of mothers of healthy, term-born infants, 10% of mothers reported clinically significant depression symptoms which is similar to population estimates (47). However, rates of perinatal depression are as high as 47% in mothers of infants born preterm (48). Study findings may, therefore, not generalize to mothers with clinical depression/anxiety or infants born preterm.

In this study of healthy term-born neonates with dMRI data collected at birth, prenatal exposure to Social Disadvantage, but not Psychosocial Stress, was independently associated with white matter microstructure at birth. However, among lower-to-higher SES infants, prenatal exposure to Psychosocial Stress was more strongly associated with altered microstructure in the inferior CB. Taken together, findings suggest that prenatal adversity across multiple socioeconomic and psychosocial domains is an important antecedent of aberrant fronto-limbic white matter fiber development at birth. Poverty reduction interventions and increased access to welfare programs that address psychosocial stressors for expectant parents may be an important target to protect fetal brain development.

## Data Availability

De-identified data is being deposited in the NIMH Data Archive and will be available after the conclusion of the eLABE 3-year wave.

