## Supplementary Information for "Exposure to Prenatal Social Disadvantage and Maternal Psychosocial Stress: Relationships to Neonatal White Matter Connectivity"

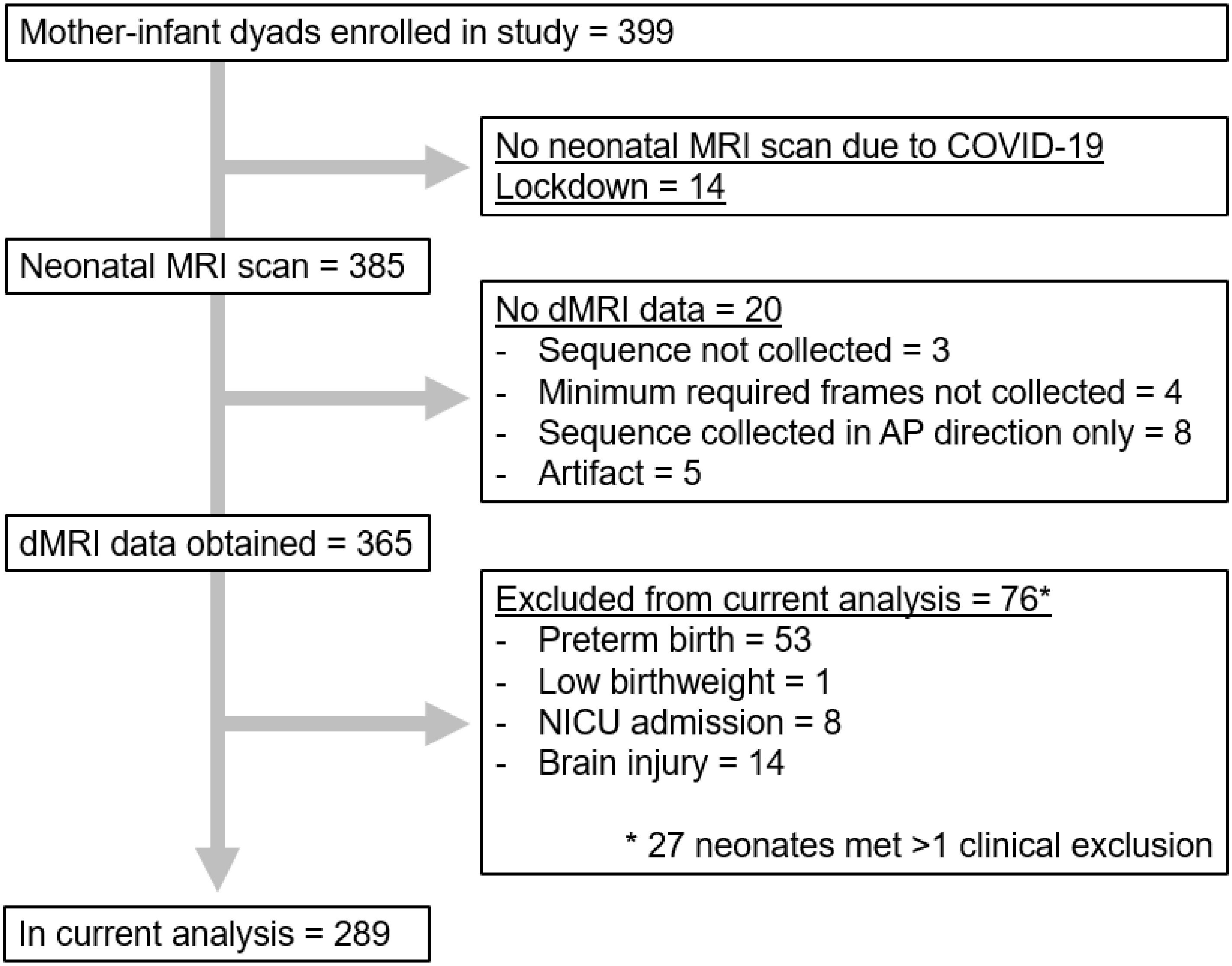


**Figure S1.** Participant flow diagram from study enrollment (*n*=399) through to inclusion in final reported data analysis (*n*=289).
dMRI, diffusion magnetic resonance imaging; AP, Anterior-to-Posterior direction; NICU, Neonatal Intensive Care Unit

**Structural Equation Modelling (SEM) of Social Disadvantage and Psychosocial Stress.**
As described in Luby et al. (1), SEM was performed in MPLUS (version 8.4) to group the observed prenatal adversity variables into a maternal Social Advantage latent factor (Income-to-Needs Ratio, Area Deprivation Index, health insurance status, highest level of education, and Healthy Eating Index) and a Psychosocial Stress latent factor (depression symptoms, perceived stress, racial discrimination, and lifetime measures of trauma and life events) (Figure S2). The SEM was performed using maximum likelihood estimation with robust standard errors to create latent factor scores for all 399 mothers, including those with partial data on observed variables. As described in Luby et al., a two factor model provided the best fit of the data. Additionally, there were low correlations of the observed variables for one factor (*e.g.*, Social Advantage) with the other factor (*e.g.,* Psychosocial Stress), supporting the grouping of the observed variables. Note that for the purposes of the current study, standardized Social Advantage values were reverse scored and termed Social Disadvantage to (a) correspond with the direction of Psychosocial Stress (higher z-scores = greater adversity) and (b) allow for easier interpretation of associations with MD and FA, which are typically inversely related in neonates (2). In the current study sample ($n$=289), Social Disadvantage and Psychosocial Stress were positively correlated (Pearson $r$ = .40, $p$ <.001).

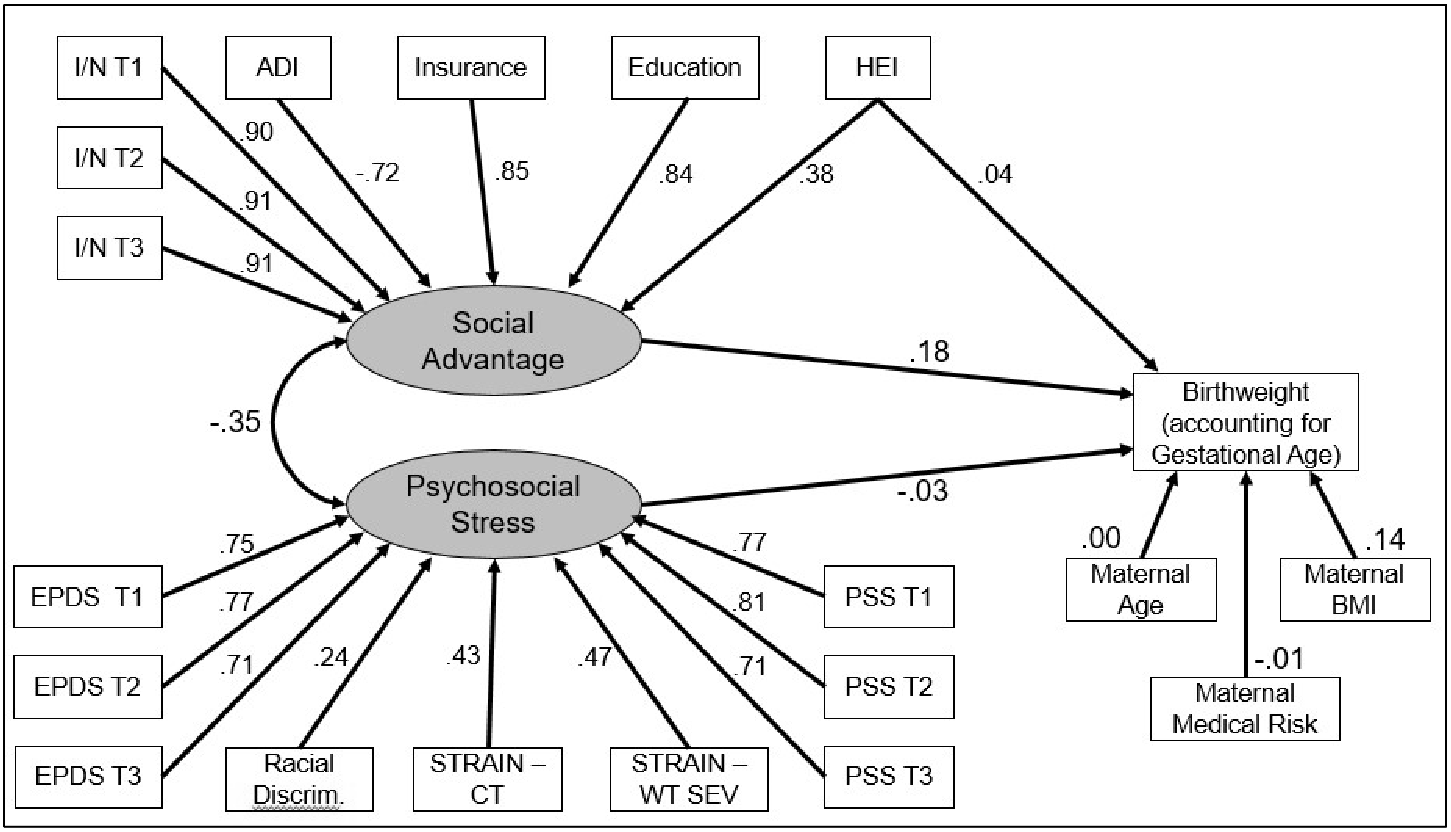


**Figure S2.** SEM illustrating the latent prenatal Social Advantage and Psychosocial Stress factors and their observed components as reported by Luby et al. (1) ($n$=399). Standardized estimates between observed and latent variables are shown. Estimates between observed variables are not shown.
T1, Trimester 1; T2, Trimester 2; T3, Trimester 3; I/N, Income-to-Needs Ratio; ADI, Area Deprivation Index; HEI, Healthy Eating Index; EPDS, Edinburgh Postnatal Depression Scale; Scale Discrim., Discrimination, STRAIN CT/WT SEV, Stress and Adversity Inventory for Adults Count/Weighted Severity; PSS, Perceived Stress Scale; BMI, Body Mass Index.

**Table S1.** Summary of association between Social Disadvantage and mean diffusivity (MD), radial diffusivity (RD), axial diffusivity (AD), and fractional anisotropy (FA) (*n*=289).

| | MD | RD | AD | FA |
|---|---|---|---|---|
| **Right Dorsal Cingulum** | | | | |
| Social Disadvantage | **-.162 (.05)**** | **-.181 (.05)**** | -.083 (.05) | .152 (.06)** |
| **Left Dorsal Cingulum** | | | | |
| Social Disadvantage | **-.127 (.05)*** | **-.145 (.05)**** | -.049 (.06) | .125 (.06)* |
| **Right Inferior Cingulum** | | | | |
| Social Disadvantage | **-.244 (.05)***** | **-.212 (.05)***** | **-.186 (.05)**** | .056 (.06) |
| **Left Inferior Cingulum** | | | | |
| Social Disadvantage | **-.234 (.05)***** | **-.208 (.05)***** | **-.204 (.05)***** | .034 (.06) |
| **Right Uncinate** | | | | |
| Social Disadvantage | **-.115 (.05)*** | -.096 (.05) | -.120 (.06)* | -.002 (.05) |
| **Left Uncinate** | | | | |
| Social Disadvantage | **-.171 (.06)**** | **-.114 (.05)*** | **-.135 (.06)*** | .049 (.05) |
| **Right Fornix** | | | | |
| Social Disadvantage | **-.129 (.05)**** | **-.149 (.05)**** | -.079 (.06) | .113 (.06)* |
| **Left Fornix** | | | | |
| Social Disadvantage | -.091 (.05) | **-.122 (.05)*** | -.020 (.05) | .122 (.06)* |

*Note*. All models adjusted for sex and postmenstrual age at scan. Standardized regression coefficients and standard errors shown.

* $p$ <.05, ** $p$ <.01, *** $p$ <.001

**Bold** values indicate significant results ($q$ <.05) after multiple comparison correction using Benjamini-Hochberg False Discovery Rate procedure.

**Table S2.** Summary of association between maternal Psychosocial Stress and mean diffusivity (MD), radial diffusivity (RD), axial diffusivity (AD), and fractional anisotropy (FA) (*n*=289).

| | MD | RD | AD | FA |
|---|---|---|---|---|
| **Right Dorsal Cingulum** | | | | |
| Psychosocial Stress | -.051 (.05) | -.041 (.05) | -.059 (.05) | .002 (.06) |
| **Left Dorsal Cingulum** | | | | |
| Psychosocial Stress | -.100 (.05) | -.086 (.05) | -.094 (.05) | .028 (.06) |
| **Right Inferior Cingulum** | | | | |
| Psychosocial Stress | -.093 (.05) | -.082 (.05) | -.062 (.06) | .040 (.06) |
| **Left Inferior Cingulum** | | | | |
| Psychosocial Stress | **-.134 (.05)**** | **-.150 (.05)**** | -.064 (.06) | .099 (.06) |
| **Right Uncinate** | | | | |
| Psychosocial Stress | -.056 (.05) | -.033 (.05) | -.068 (.06) | -.010 (.05) |
| **Left Uncinate** | | | | |
| Psychosocial Stress | -.071 (.06) | -.063 (.05) | -.017 (.06) | .088 (.05) |
| **Right Fornix** | | | | |
| Psychosocial Stress | -.026 (.05) | -.027 (.05) | -.045 (.05) | -.011 (.06) |
| **Left Fornix** | | | | |
| Psychosocial Stress | -.025 (.05) | -.049 (.05) | .021 (.05) | .081 (.06) |

*Note*. All models adjusted for sex and postmenstrual age at scan. Standardized regression coefficients and standard errors shown.
* $p$ <.05, ** $p$ <.01, *** $p$ <.001
**Bold** values indicate significant results ($q$ <.05) after multiple comparison correction using Benjamini-Hochberg False Discovery Rate procedure.

**Table S3.** Full results of hierarchical regression models linking prenatal adversity constructs with neonatal mean diffusivity (*n*=289).

| | Step 1 | | | Step 2 | | | | Change Statistics | |
|---|---|---|---|---|---|---|---|---|---|
| | **β** | **SE** | ***p*** | **β** | **SE** | ***p*** | ***q*** | **Δ $R^2$** | ***p*** |
| **Right Dorsal Cingulum** | $R^2$=.23, *p* <.001 | | | $R^2$=.25, *p* <.001 | | | | | |
| Sex | -.028 | .052 | .58 | -.022 | .051 | .66 | .87 | | |
| PMA at scan | -.478 | .052 | <.001 | -.504 | .052 | <.001 | **<.001** | | |
| Social Disadvantage | - | - | - | -.168 | .056 | .003 | **.008** | .03 | .007 |
| Psychosocial Stress | - | - | - | .015 | .055 | .79 | .94 | | |
| **Left Dorsal Cingulum** | $R^2$=.21, *p* <.001 | | | $R^2$=.22, *p* <.001 | | | | | |
| Sex | -.029 | .052 | .59 | -.022 | .052 | .67 | .87 | | |
| PMA at scan | -.454 | .053 | <.001 | -.471 | .053 | <.001 | **<.001** | | |
| Social Disadvantage | - | - | - | -.103 | .057 | .07 | .07 | .02 | .03 |
| Psychosocial Stress | - | - | - | -.059 | .056 | .29 | .94 | | |
| **Right Inferior Cingulum** | $R^2$=.23, *p* <.001 | | | $R^2$=.29, *p* <.001 | | | | | |
| Sex | -.114 | .052 | .03 | -.104 | .051 | .04 | .16 | | |
| PMA at scan | -.478 | .053 | <.001 | -.516 | .052 | <.001 | **<.001** | | |
| Social Disadvantage | - | - | - | -.246 | .055 | <.001 | **<.001** | .06 | <.001 |
| Psychosocial Stress | - | - | - | .004 | .055 | .94 | .94 | | |
| **Left Inferior Cingulum** | $R^2$=.31, *p* <.001 | | | $R^2$=.36, *p* <.001 | | | | | |
| Sex | -.119 | .050 | .02 | -.109 | .048 | .02 | .16 | | |
| PMA at scan | -.555 | .050 | <.001 | -.588 | .049 | <.001 | **<.001** | | |
| Social Disadvantage | - | - | - | -.214 | .052 | <.001 | **<.001** | .06 | <.001 |
| Psychosocial Stress | - | - | - | -.050 | .052 | .33 | .94 | | |
| **Right Uncinate** | $R^2$=.20, *p* <.001 | | | $R^2$=.21, *p* <.001 | | | | | |
| Sex | -.060 | .053 | .26 | -.056 | .053 | .30 | .69 | | |
| PMA at scan | -.446 | .054 | <.001 | -.464 | .054 | <.001 | **<.001** | | |
| Social Disadvantage | - | - | - | -.110 | .058 | .06 | .07 | .01 | .10 |
| Psychosocial Stress | - | - | - | -.012 | .057 | .84 | .94 | | |
| **Left Uncinate** | $R^2$=.13, *p* <.001 | | | $R^2$=.16, *p* <.001 | | | | | |
| Sex | -.060 | .055 | .28 | -.051 | .055 | .35 | .69 | | |
| PMA at scan | -.368 | .056 | <.001 | -.396 | .056 | <.001 | **<.001** | | |
| Social Disadvantage | - | - | - | -.169 | .060 | .005 | **.01** | .03 | .009 |
| Psychosocial Stress | - | - | - | -.006 | .059 | .91 | .94 | | |
| **Right Fornix** | $R^2$=.30, *p* <.001 | | | $R^2$=.32, *p* <.001 | | | | | |
| Sex | .003 | .050 | .95 | .008 | .049 | .87 | .87 | | |
| PMA at scan | -.547 | .050 | <.001 | -.569 | .050 | <.001 | **<.001** | | |
| Social Disadvantage | - | - | - | -.141 | .054 | .009 | **.01** | .02 | .03 |
| Psychosocial Stress | - | - | - | .029 | .053 | .59 | .94 | | |
| **Left Fornix** | $R^2$=.30, *p* <.001 | | | $R^2$=.31, *p* <.001 | | | | | |
| Sex | .007 | .049 | .89 | .010 | .049 | .84 | .87 | | |
| PMA at scan | -.549 | .050 | <.001 | -.564 | .050 | <.001 | **<.001** | | |
| Social Disadvantage | - | - | - | -.096 | .054 | .07 | .07 | .01 | .18 |
| Psychosocial Stress | - | - | - | .013 | .053 | .80 | .94 | | |

*Note.* SE, Standard Error; *q,* significance value adjusted for multiple comparisons using Benjamini-Hochberg False Discovery Rate procedure; PMA, postmenstrual age at scan
**Bold** values indicate significant results (*q* <.05) after multiple comparison correction using Benjamini-Hochberg False Discovery Rate procedure.

**Table S4.** Full results of hierarchical regression linking prenatal adversity constructs with neonatal fractional anisotropy (*n*=289).

| | Step 1 | | | Step 2 | | | | Change Statistics | |
|---|---|---|---|---|---|---|---|---|---|
| | **β** | **SE** | ***p*** | **β** | **SE** | ***p*** | ***q*** | **Δ $R^2$** | ***p*** |
| **Right Dorsal Cingulum** | $R^2$=.10, *p* <.001 | | | $R^2$=.13, *p* <.001 | | | | | |
| Sex | -.106 | .056 | .06 | -.111 | .056 | .05 | .13 | | |
| PMA at scan | .293 | .057 | <.001 | .321 | .057 | <.001 | **<.001** | | |
| Social Disadvantage | - | - | - | .179 | .061 | .003 | **.02** | .03 | .01 |
| Psychosocial Stress | - | - | - | -.068 | .060 | .26 | .57 | | |
| **Left Dorsal Cingulum** | $R^2$=.11, *p* <.001 | | | $R^2$=.13, *p* <.001 | | | | | |
| Sex | -.156 | .056 | .006 | -.160 | .056 | .004 | **.03** | | |
| PMA at scan | .280 | .056 | <.001 | .301 | .057 | <.001 | **<.001** | | |
| Social Disadvantage | - | - | - | .135 | .061 | .03 | .07 | .02 | .08 |
| Psychosocial Stress | - | - | - | -.025 | .060 | .68 | .83 | | |
| **Right Inferior Cingulum** | $R^2$=.11, *p* <.001 | | | $R^2$=.11, *p* <.001 | | | | | |
| Sex | .059 | .056 | .29 | .057 | .056 | .31 | 38 | | |
| PMA at scan | .334 | .056 | <.001 | .342 | .057 | <.001 | **<.001** | | |
| Social Disadvantage | - | - | - | .047 | .061 | .44 | .71 | <.01 | .57 |
| Psychosocial Stress | - | - | - | .021 | .060 | .72 | .83 | | |
| **Left Inferior Cingulum** | $R^2$=.10, *p* <.001 | | | $R^2$=.11, *p* <.001 | | | | | |
| Sex | -.052 | .057 | .36 | -.055 | .057 | .34 | .38 | | |
| PMA at scan | .305 | .057 | <.001 | .305 | .058 | <.001 | **<.001** | | |
| Social Disadvantage | - | - | - | -.006 | .062 | .92 | .97 | .01 | .22 |
| Psychosocial Stress | - | - | - | .101 | .061 | .10 | .57 | | |
| **Right Uncinate** | $R^2$=.26, *p* <.001 | | | $R^2$=.26, *p* <.001 | | | | | |
| Sex | -.114 | .051 | .03 | -.114 | .051 | .03 | .11 | | |
| PMA at scan | .491 | .051 | <.001 | .492 | .052 | <.001 | **<.001** | | |
| Social Disadvantage | - | - | - | .002 | .056 | .97 | .97 | <.00 | .98 |
| Psychosocial Stress | - | - | - | -.011 | .055 | .84 | .84 | | |
| **Left Uncinate** | $R^2$=.18, *p* <.001 | | | $R^2$=.19, *p* <.001 | | | | | |
| Sex | -.004 | .053 | .99 | -.004 | .053 | .94 | .94 | | |
| PMA at scan | .426 | .054 | <.001 | .429 | .054 | <.001 | **<.001** | | |
| Social Disadvantage | - | - | - | .016 | .058 | .78 | .97 | .01 | .24 |
| Psychosocial Stress | - | - | - | .082 | .058 | .16 | .57 | | |
| **Right Fornix** | $R^2$=.09, *p* <.001 | | | $R^2$=.11, *p* <.001 | | | | | |
| Sex | -.081 | .057 | .154 | -.084 | .057 | .14 | .22 | | |
| PMA at scan | .289 | .057 | <.001 | .311 | .058 | <.001 | **<.001** | | |
| Social Disadvantage | - | - | - | .140 | .062 | .03 | .07 | .02 | .08 |
| Psychosocial Stress | - | - | - | -.066 | .061 | .28 | .57 | | |
| **Left Fornix** | $R^2$=.08, *p* <.001 | | | $R^2$=.10, *p* <.001 | | | | | |
| Sex | -.091 | .057 | .11 | -.097 | .057 | .09 | .18 | | |
| PMA at scan | .261 | .057 | <.001 | .278 | .058 | <.001 | **<.001** | | |
| Social Disadvantage | - | - | - | .107 | .062 | .09 | .17 | .02 | .08 |
| Psychosocial Stress | - | - | - | .039 | .061 | .52 | .83 | | |

*Note.* SE, Standard Error; *q,* significance value adjusted for multiple comparisons using Benjamini-Hochberg False Discovery Rate procedure; PMA, postmenstrual age at scan
**Bold** values indicate significant results (*q* <.05) after multiple comparison correction using Benjamini-Hochberg False Discovery Rate procedure.

**The Corpus Callosum (CC) as a Negative Control Tract.**

In line with prior work from our group (3), the CC was selected as a negative control tract because, like the cingulum bundle (CB), the CC is long-range tract with multiple branching fibers and it has a similar anterior-posterior orientation (4). However, unlike the CB, uncinate, and fornix, the CC does not connect limbic system structures (*i.e.,* amygdala and hippocampus) with the frontal cortex (5). We performed multivariable regressions including Social Disadvantage, Psychosocial Stress, sex, and infant PMA at scan as independent variables fitted to CC MD and FA as dependent variables. MA and FA were extracted from the CC using identical methods as the CB, uncinate, and fornix. As shown in Table S5, neither prenatal exposure to Social Disadvantage nor Psychosocial Stress were associated with MD or FA in the CC ($p$>.05).

**Table S5.** Associations between prenatal adversity constructs and neonatal mean diffusivity (MD) fractional anisotropy (FA) in the corpus callosum (*n*=289).

| | **β** | **SE** | ***p*** |
|---|---|---|---|
| **Corpus Callosum MD** | | $R^2$=.30, *p* <.001 | |
| Sex | -.005 | .049 | .91 |
| PMA at scan | -.555 | .050 | **<.001** |
| Social Disadvantage | -.039 | .054 | .47 |
| Psychosocial Stress | -.035 | .053 | .51 |
| **Corpus Callosum FA** | | | |
| Sex | -.094 | .053 | .08 |
| PMA at scan | .426 | .054 | **<.001** |
| Social Disadvantage | -.090 | .058 | .12 |
| Psychosocial Stress | -.041 | .057 | .47 |

*Note.* SE, Standard Error; PMA, postmenstrual age at scan

**Psychosocial Stress in Extremely Low and Lower-to-Higher Socioeconomic Status (SES) Groups.** Because a bivariate association was observed between Psychosocial Stress and MD in the left inferior CB prior to accounting for broad Social Disadvantage (Table S2), we examined whether the strength of the association between maternal Psychosocial Distress and inferior CB connectivity varied as a function of family SES. To create family SES groups for moderation analysis, the sample was dichotomized using mean Income-to-Needs Ratio (INR) across trimesters. INR was selected to dichotomize the sample because INR loaded most heavily on the latent Social Disadvantage construct (1) (Figure S2). In line with the large, prospective Fragile Families and Child Wellbeing Study and other reports (6–8), INR values below 200% of the national poverty threshold were categorized as extremely low family SES (*n*=179), and values at or above 200% of the national poverty threshold categorized as lower-to-higher family SES (*n*=105). INR values were missing for five mothers. As expected, mothers in the extremely low SES group had higher Psychosocial Stress scores (*m*= 0.68, SD= 1.02) than mothers in the lower-to-higher SES group (*m*= -0.46, SD= 0.80, *t*= -6.78, *p*<.001).

To test the interaction between Psychosocial Stress and family SES group on MD in the left inferior CB, moderation analysis was performed using the PROCESS procedure for SPSS (9). Family SES group and Psychosocial Stress were included as main effects, along with a mean-centered interaction term and covariate factors (sex and infant PMA at scan). The interaction between Psychosocial Stress and family SES group was significant (*p* =.008), such that the association between Psychosocial Stress and MD in the left inferior CB was stronger in the lower-to-higher SES group than in the extremely low SES group (see also Figure 2, Main Text).

**Table S6.** Moderation analysis of the interaction between family SES group and Psychosocial stress (*n*=284).

| | B | SE | *p* |
|---|---|---|---|
| **Left Inferior Cingulum MD** | | $R^2$=.38, *p* <.001 | |
| Sex | -.123 | .048 | **.01** |
| PMA at scan | -.579 | .048 | **<.001** |
| Family SES group | .319 | .111 | **.004** |
| Psychosocial Stress | -.102 | .052 | .05 |
| Interaction: Family SES group x Psychosocial Stress | -.305 | 115 | **.008** |

*Note.* Unstandardized coefficients (B) from PROCESS output shown. SE, Standard Error; PMA, postmenstrual age at scan

For completeness, we also ran a multivariable regression model among the lower-to-higher SES group to examine whether the association between Psychosocial Stress and MD in the left inferior CB persisted after also accounting for individual differences in social background. As shown in Table S7, Psychosocial Stress remained significant (*p* =.006) among the lower-to-higher SES group even after accounting for Social Disadvantage factor scores, which were not significant (*p* =.67).

**Table S7.** Associations between prenatal adversity constructs and neonatal mean diffusivity (MD) fractional anisotropy (FA) in family SES groups (*n*=284).

| | **Extremely Low SES (*n*=179)** | | | **Lower-to-Higher SES (*n*=105)** | | |
|---|---|---|---|---|---|---|
| | **β** | **SE** | ***p*** | **β** | **SE** | ***p*** |
| **Left Inferior Cingulum MD** | $R^2$=.36, *p* <.001 | | | $R^2$=.41, *p* <.001 | | |
| Sex | -.167 | .063 | **.009** | -.060 | .073 | .41 |
| PMA at scan | -.570 | .058 | **<.001** | -.623 | .087 | **<.001** |
| Social Disadvantage | -.283 | .179 | .12 | -.057 | .130 | .67 |
| Psychosocial Stress | .026 | .062 | .67 | -.268 | .096 | **.006** |

*Note.* SE, Standard Error; PMA, postmenstrual age at scan

**Confounding Factors: Supplemental Analysis Addressing Maternal Medical Risk (MMR) in Pregnancy.** Supplemental analysis was performed to account for the potentially confounding role of maternal medical co-morbidities during pregnancy on offspring white matter connectivity at birth. A MMR index was calculated for each mother using questionnaire data and chart review (1). This validated MMR index is a weighted sum of maternal morbidities including advanced age, pre-gestational diabetes, placenta previa, asthma, hypertension, prior C-section delivery, pre-eclampsia, cardiac disease, renal disease, sickle cell disease, lupus, and human immunodeficiency virus (10–12). Higher MMR index scores indicate increased medical risk. Mothers in this study were relatively healthy with an overall mean MMR of 1.01 (SD= 1.26, range: 0 – 8).

Non-parametric spearman's rho correlations were used to screen for associations between the MMR index and offspring white matter tract MD and FA at birth. MMR index was only found to be correlated with MD in the left dorsal CB at birth (ρ=.13, *p*=.03); there were no other associations for any other white matter tracts (null results available in full upon request). Also of note, MMR index was not associated with either Social Disadvantage (ρ=-.01, *p*=.84) or Psychosocial Stress (ρ=-.07, *p*=.23) scores.

The regression model for MD in the left dorsal CB was re-run including MMR index as a covariate (Table S8). After accounting for infant PMA at scan (*p*<.001), MMR was no longer significant (*p*=.49). The finding concerning Social Disadvantage was unchanged (*p*>.05, compare with Table S3).

**Table S8.** Maternal Medical Risk, prenatal adversity, and mean diffusivity (MD) in the left dorsal portion of the cingulum bundle (*n*=289).

| | **β** | **SE** | ***p*** |
|---|---|---|---|
| **Left Dorsal Cingulum MD** | | $R^2$=.23, *p* <.001 | |
| Sex | -.023 | .052 | .66 |
| PMA at scan | -.468 | .053 | **<.001** |
| MMR index | .029 | .041 | .49 |
| Social Disadvantage | -.102 | .057 | .07 |
| Psychosocial Stress | -.057 | .056 | .31 |

*Note.* SE, Standard Error; PMA, postmenstrual age at scan

**Confounding Factors: Supplemental Analysis Addressing Prenatal Cannabis and Tobacco Exposure.** Supplemental analysis was performed to account for the potentially confounding role of prenatal cannabis and tobacco exposure on white matter connectivity at birth. During pregnancy, mothers completed self-report surveys detailing the frequency of cannabis and tobacco use per trimester. When available (42.6% of the current sample), cannabis exposure information was supplemented with maternal urine drug screen (UDS) performed at the discretion of the treating physician as part of obstetric care and recorded in patient medical records. To combine self-report cannabis data with UDS positive for tetrahydrocannabinol metabolites, frequency of self-reported cannabis use was coded never = 0 and all other responses (daily, weekly but not every day, monthly but not every week) = 1. For comparability, self-report tobacco use was also binarized and coded as no use = 0 versus one or more cigarettes per day = 1. Seventy-seven (27%) mothers reported cannabis use and/or had a positive UDS. Thirty-nine (14%) mothers reported cigarette use. Independent samples *t*-tests indicated that mothers who reported cannabis use and/or had a positive UDS had higher levels of Social Disadvantage ($p$<.001) and Psychosocial Stress ($p$<.001) than mothers who had no cannabis exposure (Table S9). Similarly, mothers who reported tobacco use had higher levels of Social Disadvantage ($p$<.001) and Psychosocial Stress ($p$<.001) than mothers who reported no tobacco use.

**Table S9.** Comparison of prenatal adversity factors between drug exposure groups (*n*=289).

| | **Tobacco Exposure (*n* = 39)** | **No Tobacco Exposure (*n* = 250)** | ***t*** | ***p*** |
|---|---|---|---|---|
| Social Disadvantage, *m* (SD) | 0.79 (0.43) | -0.11 (1.01) | -9.62 [a] | <.001 |
| Psychosocial Stress, *m* (SD) | 0.51 (0.99) | -0.08 (0.98) | -3.46 | <.001 |
| | **Cannabis Exposure (*n* = 77)** | **No Cannabis Exposure (*n* = 212)** | ***t*** | ***p*** |
| Social Disadvantage, *m* (SD) | 0.72 (0.34) | -0.25 (1.04) | -11.85 [a] | <.001 |
| Psychosocial Stress, *m* (SD) | 0.38 (0.93) | -0.14 (0.99) | -4.00 | <.001 |

[a] *t* statistic and corresponding *p*-value corrected for unequal variances between groups

Independent samples *t*-tests were used to screen for differences in neonatal white matter tract MD and FA at birth between drug exposure groups. Prenatal cannabis exposure was associated with lower MD in the inferior portion of the left CB ($t$ (287) = 2.06, $p$=.04) and higher FA in the left fornix ($t$ (287) = -2.37, $p$=.02). Cannabis exposure was not associated with any other white matter tracts. Prenatal tobacco exposure was not associated with MD or FA in any white matter tracts (null results available in full upon request). Regression models for left inferior CB MD and left fornix FA were re-run including binarized cannabis exposure as a covariate (Table S10). After accounting for infant PMA at scan ($p$<.001), prenatal cannabis exposure was no longer significant ($p$ >.05). Key study findings concerning Social Disadvantage were unchanged (see Tables S3 and S4).

**Table S10.** Prenatal cannabis exposure, prenatal adversity, and white matter at birth (*n*=289).

| | **β** | **SE** | ***p*** |
|---|---|---|---|
| **Left Inferior Cingulum MD** | | $R^2$=.36, $p$ <.001 | |
| Sex | -.110 | .048 | **.02** |
| PMA at scan | -.590 | .049 | **<.001** |
| Prenatal cannabis exposure | .015 | .052 | .78 |
| Social Disadvantage | -.221 | .057 | **<.001** |
| Psychosocial Stress | -.051 | .052 | .33 |
| **Left Fornix FA** | | $R^2$=.10, $p$ <.001 | |
| Sex | -.102 | .057 | .07 |
| PMA at scan | .267 | .058 | **<.001** |
| Prenatal cannabis exposure | .098 | .062 | .12 |
| Social Disadvantage | .066 | .067 | .33 |
| Psychosocial Stress | .033 | .061 | .59 |

*Note.* SE, Standard Error; PMA, postmenstrual age at scan
